# EFFECT OF YOGA-BASED INTERVENTION ON STRESS AMONG UNIVERSITY STUDENTS: A RANDOMIZED CONTROL TRIAL

**DOI:** 10.1101/2025.11.27.25340139

**Authors:** Sai Charan Reddy Bhimavarapu, Akanksha Rathore, Kanika Ahlawat, KM. Jaya Rai, Nandini Rastogi, Vaseem Saifi

## Abstract

University students are particularly vulnerable to stress due to academic demands, social pressures, and transitional life challenges. Chronic stress among this population can adversely affect academic performance, mental health, and overall well-being. Yoga, a holistic practice combining physical postures, breath control, and mindfulness, is increasingly recognized as a non-pharmacological intervention for stress reduction. However, limited evidence exists from randomized controlled trials specifically evaluating the impact of breath awareness within yoga practices among students.

**Methods:** *Introduction:* This randomized controlled trial was conducted on 60 university students aged 18–25 years with moderate to high perceived stress levels (PSS-10 score ≥14). Participants were randomly assigned to one of two groups: yoga with breath awareness (n=30) or yoga without breath awareness (n=30). Both groups underwent a structured yoga intervention consisting of loosening exercises, asanas, pranayama, and relaxation over 30 consecutive days, five days per week, with each session lasting 45 minutes. Stress levels were assessed pre- and post-intervention using the Perceived Stress Scale (PSS-10). Statistical analyses included paired and unpaired t-tests, with significance set at p<0.05.

*Results:* Both groups demonstrated significant reductions in perceived stress scores following the intervention. The yoga with breath awareness group showed a mean stress reduction of 4.43 points (19.25%, p=0.0001), while the yoga without breath awareness group showed a reduction of 4.20 points (18.42%, p=0.0026). However, inter-group comparisons revealed no statistically significant difference in stress reduction between the two groups (p=0.8730), suggesting comparable efficacy.

*Conclusion:* A 30-day structured yoga intervention effectively reduced perceived stress levels in university students, irrespective of breath awareness. Although the group practicing yoga with breath awareness exhibited slightly greater stress reduction, the difference was not statistically significant. These findings support the use of yoga as an accessible, non-pharmacological strategy for stress management in university populations. Future research with larger sample sizes, extended duration, and incorporation of physiological markers is warranted to further explore the role of breath-focused practices.

## Introduction

Stress is a common psychological and physiological response to challenging situations and is particularly prevalent among university students due to academic pressure, lifestyle changes, financial instability, and social factors^[1]^. Chronic stress is known to deregulate the hypothalamic-pituitary-adrenal (HPA) axis and the autonomic nervous system, leading to impaired cognitive functions, hormonal imbalances, and reduced emotional resilience^[2]^.Recent research estimates that nearly one-third of university students experience moderate to severe stress, which can precipitate anxiety, depression, sleep disorders, and even substance abuse^[3]^.Although pharmacological therapies such as SSRIs and SNRIs are commonly prescribed for stress-related disorders, they often come with side effects including emotional dulling, weight gain, and metabolic disturbances^[4].^ This has prompted a growing interest in non-pharmacological, holistic interventions that can address stress without adverse outcomes ^[5]^.

Yoga, a traditional mind-body practice that integrates physical postures (asanas), breath regulation (pranayama), and meditation, has shown considerable promise in promoting mental and physical well-being ^[6]^. Evidence suggests that yoga down regulates stress markers such as cortisol, improves heart rate variability, and positively modulates the autonomic nervous system^[7][8]^. The inclusion of breath awareness in yoga further enhances its therapeutic potential by shifting the balance toward parasympathetic dominance, facilitating emotional regulation and relaxation ^[9][10]^.

Several randomized controlled trials have reported the effectiveness of yoga in reducing stress, anxiety, and depressive symptoms in student and professional populations ^[11][12]^. Emerging research also indicates that conscious breath control in yoga practices leads to improved cognitive function, emotional regulation, and neuroendocrine stability compared to interventions lacking breath awareness ^[13]^.

Despite this growing body of evidence, there remains a gap in research directly comparing yoga practices with and without breath awareness among stressed university students. This study aims to explore the impact of both forms of yoga on perceived stress using the validated Perceived Stress Scale (PSS-10), with the goal of determining whether conscious breath control provides additional benefits ^[14]^.

## Materials and methods

### Ethics

The research included human participants. Prior to enrollment, all potential participants were informed about the study’s objectives, procedures, possible benefits, and any associated risks. They were assured that their participation was entirely voluntary and that they could leave the study at any point without having to give a reason. Informed written consent was obtained from those who agreed to take part. The study received ethical clearance from the Institutional Ethics Committee of Swami Vivekanand Subharti University, Meerut, Uttar Pradesh (Reference No: CTRI/2024/07/071527).

### Study design and settings

This prospective, randomized controlled trial was conducted at Maharishi Aurobindo Subharti College and Hospital of Naturopathy and Yogic Sciences, Swami Vivekanand Subharti University, Meerut. The study aimed to assess the impact of yoga-based interventions—with and without breath awareness—on perceived stress levels among university students over a 30-day period, with sessions conducted five days per week.

### Participants

A total of 104 university students from Swami Vivekanand Subharti University, Meerut, Uttar Pradesh were screened for eligibility. Inclusion criteria required participants to be between 18 to 25 years of age, enrolled in university, and experiencing moderate to high levels of stress, as measured by a Perceived Stress Scale-10 (PSS-10) score of 14 or above. Individuals with severe physical or psychological conditions, ongoing stress-related medication, or menstruation during assessment were excluded. The enrolment and allocation are shown in Figure 1.

**Fig 1:**
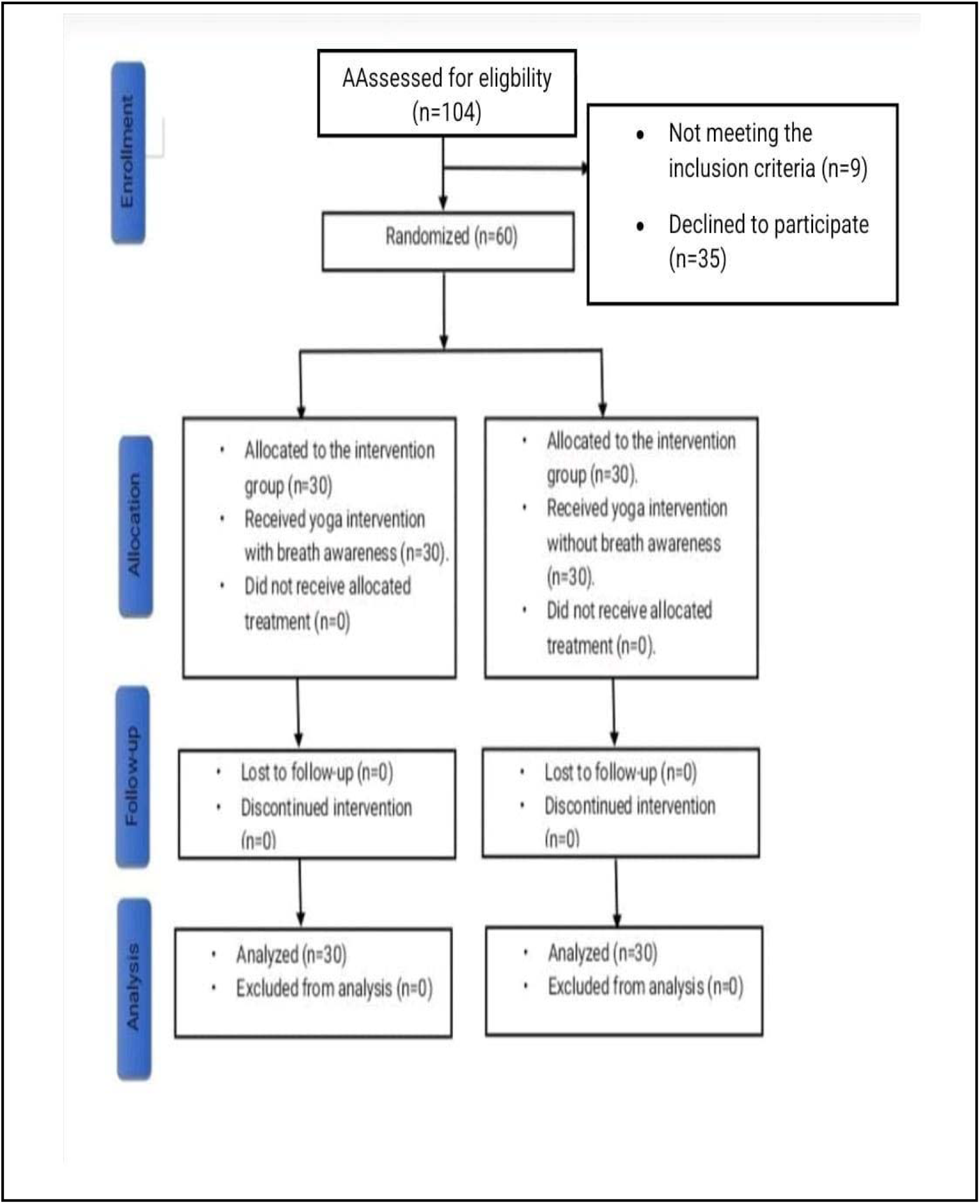
Research participant’s recruitment and their participation pattern in a flow chart.

### Intervention

Following screening, 60 eligible participants were selected through purposive sampling and randomly assigned into two groups (n=30 each) using computer-generated randomization. Group A received a yoga intervention with breath awareness, while Group B received yoga without breath awareness. All participants were informed about the nature of the study and provided written informed consent prior to participation. Confidentiality was maintained, and no participant received any additional therapies or dietary modifications during the intervention period.

The intervention spanned 30 days, with sessions conducted five days per week, each lasting 45 minutes. Participants were randomly assigned to two groups: Group A received a yoga intervention incorporating breath awareness, while Group B followed the same yoga protocol without breath awareness ^[15]^.

Each session began with a brief prayer and loosening exercises, followed by eight structured yoga postures (asanas): Trikonasana, Tiryak Tadasana, Setu Bandhasana, Pavanamuktasana, Bhujangasana, Marjariasana, Ardha Matsyendrasana, and Shashankasana ^[16]^. These asanas were practiced for approximately 25 minutes, with each posture held for 2.5 minutes. This was followed by a 10-minute pranayama segment that included Nadi Shodhana, Bhramari, and Anulom Vilom, along with three rounds of chanting ’OM’ ^[17]^. The session concluded with a short relaxation phase and a closing prayer.

**Fig 2:**
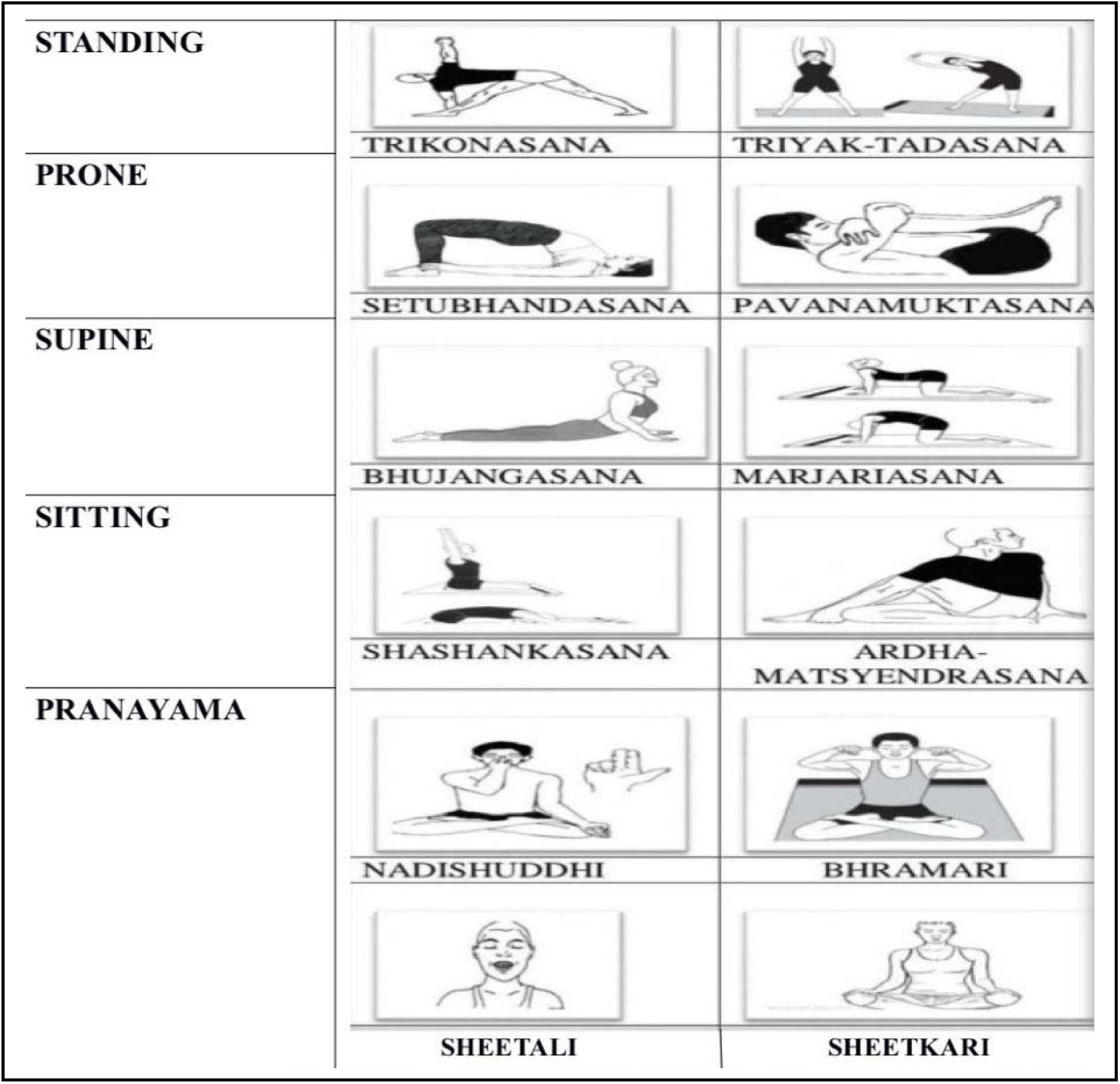
Brief procedure of the yoga protocol.

Participants in Group A were specifically instructed to maintain conscious breath awareness throughout the practice, especially during transitions between asanas and pranayama techniques ^[18]^. Group B followed the same physical sequence but without any direction regarding breath focus, concentrating only on the physical movements.

Before each session, participants were asked to maintain an empty stomach or allow a minimum two-hour gap after meals, wear loose and comfortable clothing, and avoid electronic distractions. Attendance was recorded daily, and any physical discomfort or challenges were documented. No additional lifestyle interventions or therapies were permitted during the course of the study ^[19]^.

Pre- and post-intervention assessments were conducted using the Perceived Stress Scale (PSS-10) to evaluate changes in stress level.

### Assessment

The Perceived Stress Scale-10 (PSS-10) was used as the primary assessment tool to evaluate participants’ stress levels before and after the 30-day intervention. The PSS-10 is a widely validated psychometric instrument designed to measure perceived stress, capturing how unpredictable, uncontrollable, and overloaded respondents find their lives during the past month ^[20]^. It includes 10 items scored on a five-point Likert scale ranging from 0 (never) to 4 (very often), with total scores ranging from 0 to 40; higher scores indicate greater perceived stress ^[21]^.

**Fig 3:**
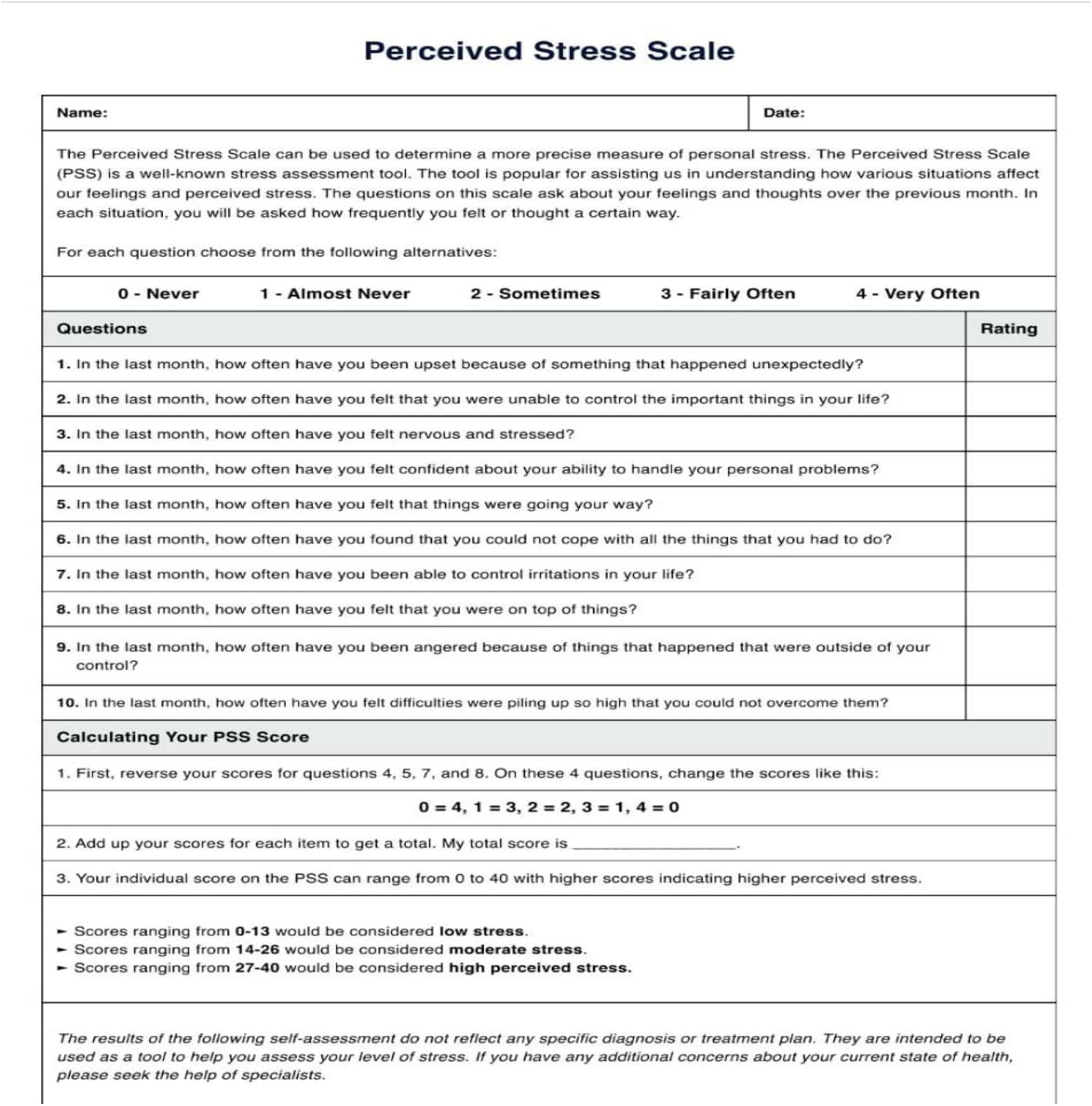
Perceived Stress Scale-10 (PSS-10)

Pre- and post-intervention assessments were conducted in a quiet, supervised environment to ensure accuracy and minimize bias. Participants completed the PSS-10 at baseline (Day 0) and again at the end of the intervention (Day 30). The same instructions and conditions were maintained during both assessments. The PSS-10 has demonstrated high internal consistency and test-retest reliability among young adults in mental health intervention studies ^[22]^.

### Statistical analysis

Data were analyzed using SPSS version 23. The normality of pre- and post-intervention PSS-10 scores in both groups was checked using the Shapiro-Wilk test. As the data followed a normal distribution, parametric tests were applied. An independent (unpaired) t-test was used to compare between-group scores, while a paired t-test assessed within-group differences. A p-value less than 0.05 was considered statistically significant for all comparisons.

## Results

A total of 104 university students were assessed for eligibility. Based on the inclusion and exclusion criteria using the Perceived Stress Scale (PSS-10), 60 participants were selected for the study. The sample size was calculated using Cohen’s formula for a moderate effect size (d = 0.5), 80% power, and 5% significance level. A paired design (pre- and post-assessment) was adopted to enhance statistical power. All selected students completed the study with no dropouts. Participants were randomly assigned into two equal groups (n = 30 each) using a computer-generated randomization sequence:

- Group A (Cases): Yoga with breath awareness
- Group B (Controls): Yoga without breath awareness

The demographic analysis showed no significant differences in age or gender between the groups, indicating proper randomization. Most participants were in the 18–21 age range.

**Table 1:** Comparison of Cases and Controls with Age.

| Age groups | Cases | % | Controls | % | Total | % |
| --- | --- | --- | --- | --- | --- | --- |
| 18-19yrs | 15 | 50.00 | 19 | 63.33 | 34 | 56.67 |
| 20-21yrs | 10 | 33.33 | 10 | 33.33 | 20 | 33.33 |
| $\geq 22$yrs | 5 | 16.67 | 1 | 3.33 | 6 | 10.00 |
| Mean | 19.73 |  | 19.47 |  | 19.60 |  |
| SD | 1.82 |  | 1.36 |  | 1.60 |  |
| Total | 30 | 100.00 | 30 | 100.00 | 60 | 100.00 |
| Chi-square=0.6010, p=0.4380 |  |  |  |  |  |  |

**Figure 4:**
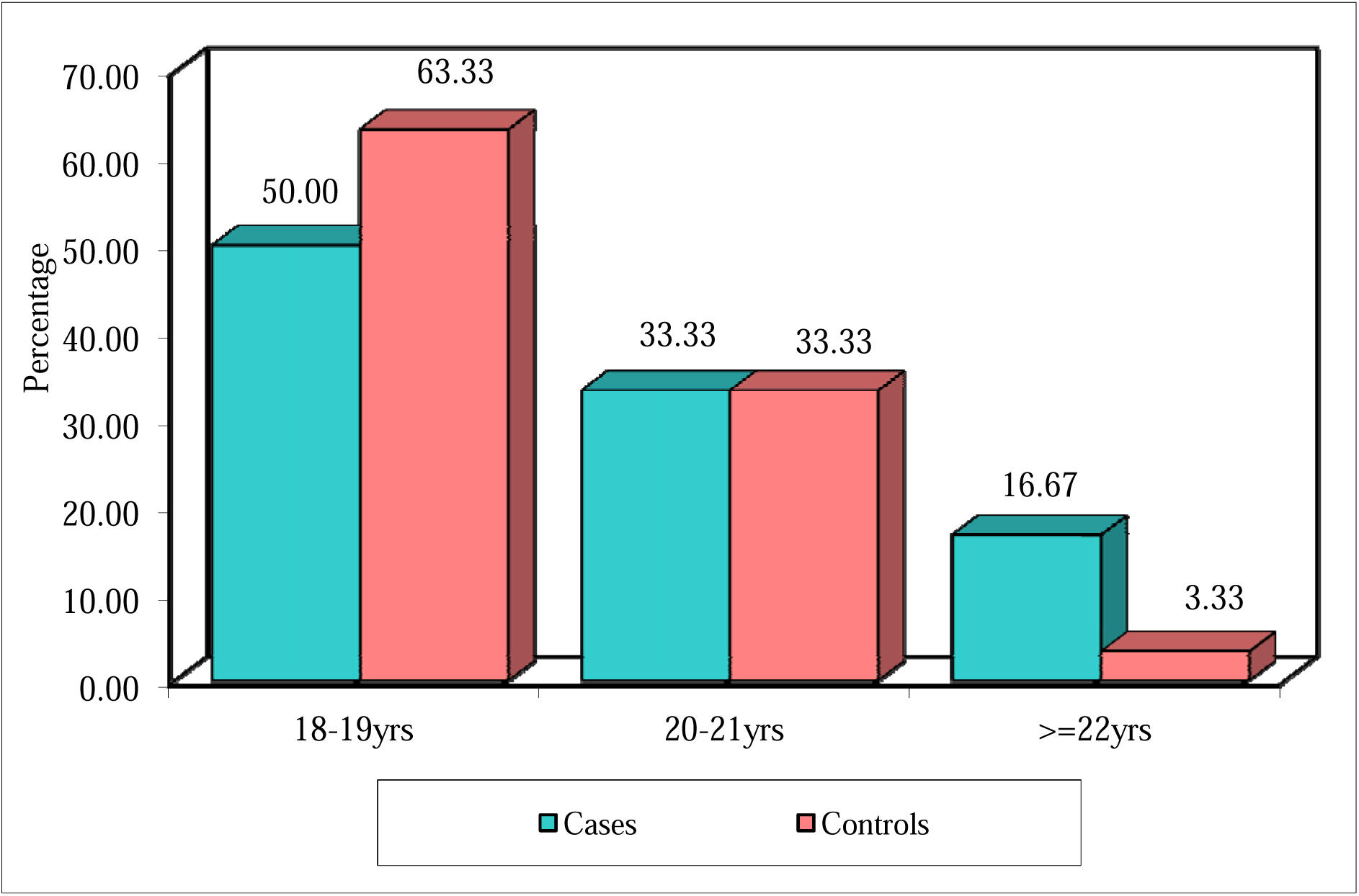
Bar Graph – Age Distribution of Cases and Controls.

**Table 2:** Comparison of Cases and Controls with Gender.

| Gender | Cases | % | Controls | % | Total | % |
| --- | --- | --- | --- | --- | --- | --- |
| Male | 14 | 46.67 | 17 | 56.67 | 31 | 51.67 |
| Female | 16 | 53.33 | 13 | 43.33 | 29 | 48.33 |
| Total | 30 | 100.00 | 30 | 100.00 | 60 | 100.00 |
| Chi-square=3.1370, p=0.2080 |  |  |  |  |  |  |

**Figure 5:**
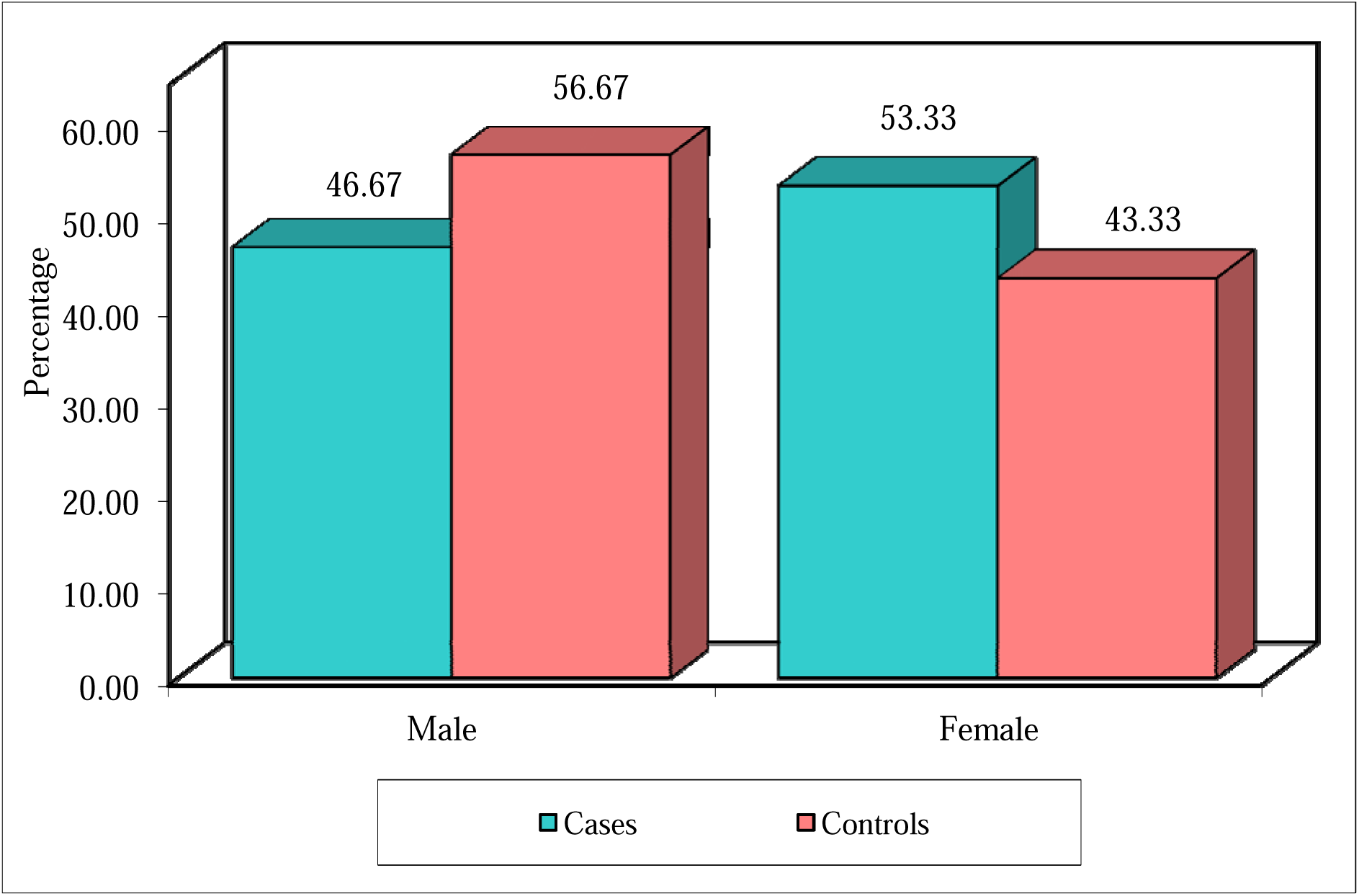
Bar Graph – Gender Distribution of Cases and Controls.

The Shapiro-Wilk test confirmed that pretest and posttest PSS scores in both groups followed a normal distribution (p > 0.05), supporting the use of parametric tests.

**Table 3:** Normality of Pretest and Posttest Stress Scores in Cases and Controls by Shapiro-Wilk Test.

| Stress at | Group | Shapiro-Wilk | df | p-value |
| --- | --- | --- | --- | --- |
| Pretest | Cases | 0.9710 | 30 | 0.5610 |
|  | Controls | 0.9280 | 30 | 0.0569 |
| Posttest | Cases | 0.9490 | 30 | 0.1550 |
|  | Controls | 0.9490 | 30 | 0.1550 |
| Difference | Cases | 0.9620 | 30 | 0.3440 |
|  | Controls | 0.9390 | 30 | 0.0860 |

At baseline, the mean PSS score was 23.03 ± 4.28 in the case group and 22.80 ± 5.87 in the control group. Post-intervention, both groups showed an identical mean score of 18.60 ± 3.20.

**Table 4:** Summary of Pretest and Posttest Stress Scores in Cases and Controls.

| Times | Group | Means | Std.Dev. | Std.Err. | Median |
| --- | --- | --- | --- | --- | --- |
| Pretest | Cases | 23.03 | 4.28 | 0.78 | 23.00 |
|  | Controls | 22.80 | 5.87 | 1.07 | 22.00 |
| Posttest | Cases | 18.60 | 3.20 | 0.58 | 18.00 |
|  | Controls | 18.60 | 3.20 | 0.58 | 18.00 |
| Difference | Cases | 4.43 | 3.81 | 0.70 | 5.00 |
|  | Controls | 4.20 | 6.98 | 1.28 | 4.00 |

A statistically significant reduction in stress was observed within each group:

- Cases: Mean reduction = 4.43 points (19.25%); t = 6.3703, p = 0.0001
- Controls: Mean reduction = 4.20 points (18.42%); t = 3.2935, p = 0.0026

**Table 5:** Comparison of Pretest and Posttest Stress Scores in Cases and Controls by Paired t-test.

| Groups | Times | Mean | SD | Mean Diff. | SD Diff. | % of effect | t-value | P-value |
| --- | --- | --- | --- | --- | --- | --- | --- | --- |
| Cases | Pretest | 23.03 | 4.28 | 4.43 | 3.81 | 19.25 | 6.3703 | 0.0001* |
|  | Posttest | 18.60 | 3.20 |  |  |  |  |  |
| Controls | Pretest | 22.80 | 5.87 | 4.20 | 6.98 | 18.42 | 3.2935 | 0.0026* |
|  | Posttest | 18.60 | 3.20 |  |  |  |  |  |

**Figure 6:**
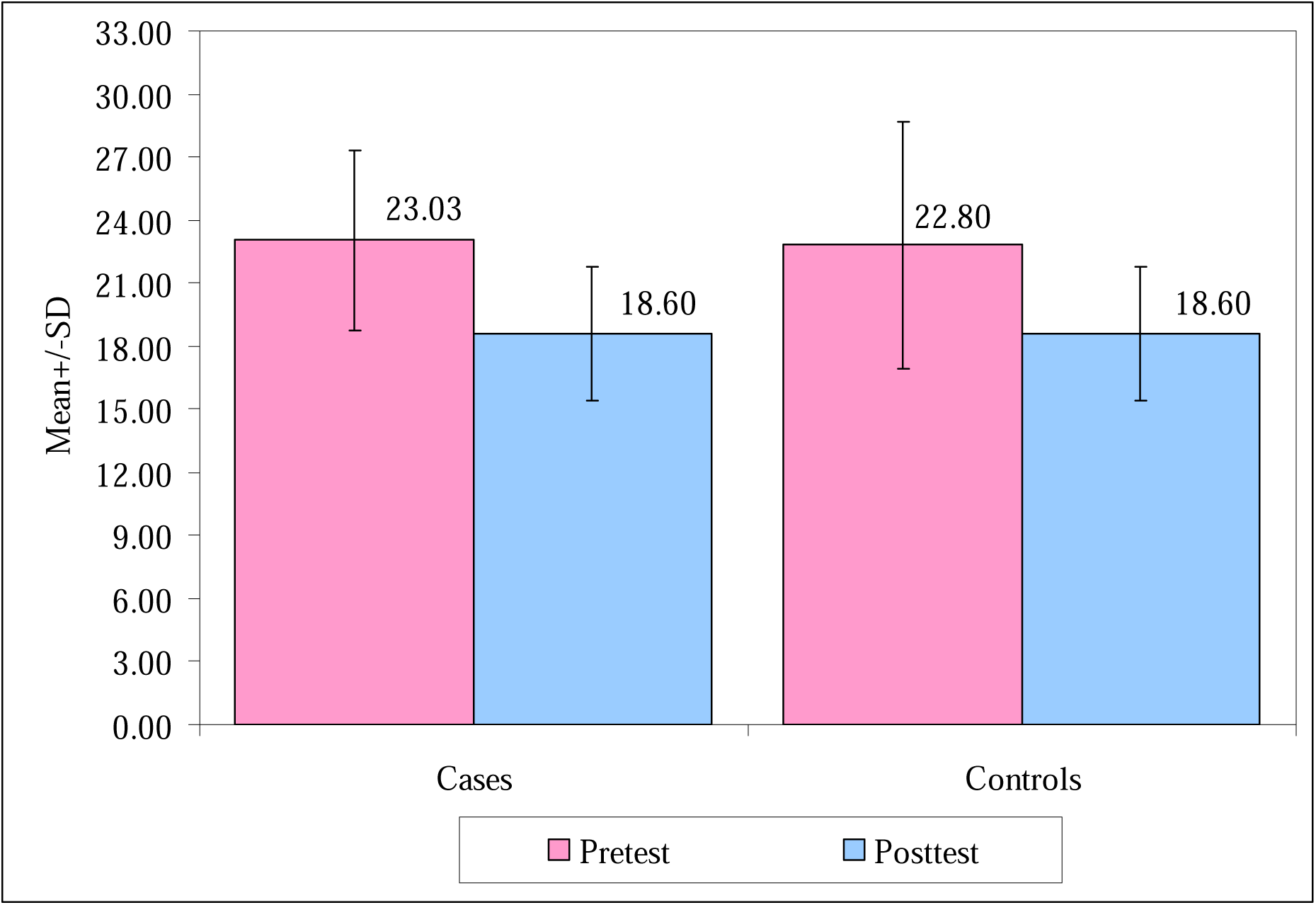
Bar Chart – Pretest and Posttest Mean Stress Scores in Cases and Controls.

No significant difference was found between the groups for pretest (p = 0.8609), posttest (p = 1.0000), or change in scores (p = 0.8730), suggesting that both interventions were equally effective in reducing perceived stress.

**Table 6:** Comparison of Pretest and Posttest Stress Scores in Cases and Controls by Unpaired t-test.

| Times | Group | n | Mean | SD | SE | t-value | P-value |
| --- | --- | --- | --- | --- | --- | --- | --- |
| Pretest | Cases | 30 | 23.03 | 4.28 | 0.78 | 0.1760 | 0.8609 |
|  | Controls | 30 | 22.80 | 5.87 | 1.07 |  |  |
| Posttest | Cases | 30 | 18.60 | 3.20 | 0.58 |  |  |
|  | Controls | 30 | 18.60 | 3.20 | 0.58 | 0.0000 | 1.0000 |
| Difference | Cases | 30 | 4.43 | 3.81 | 0.70 |  |  |
|  | Controls | 30 | 4.20 | 6.98 | 1.28 | 0.1606 | 0.8730 |

**Fig 7:**
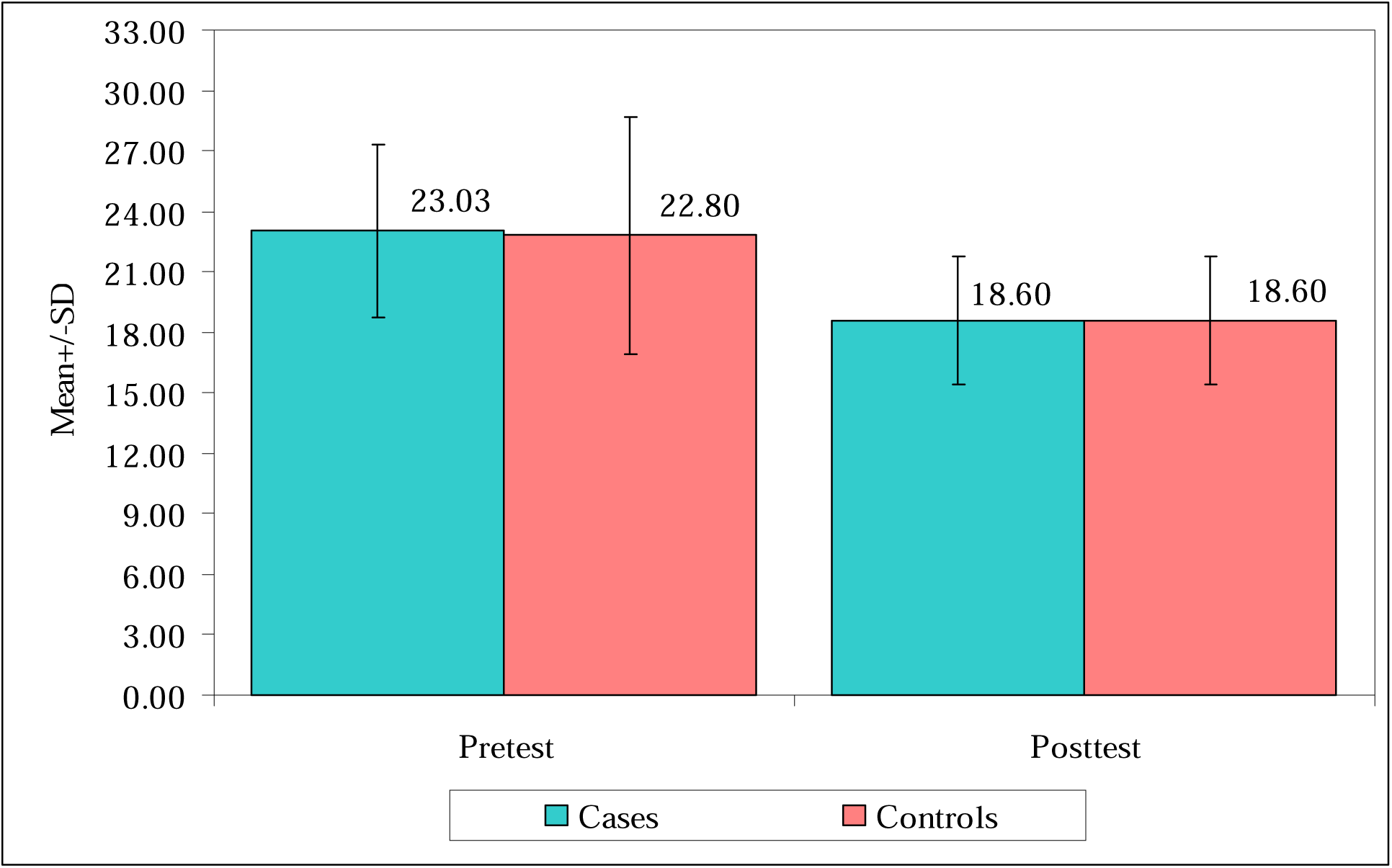
Comparisons of cases and controls with pretest and posttest stress scores.

## Discussion

This randomized controlled trial evaluated the impact of a 30-day structured yoga intervention on perceived stress among university students, focusing on the role of breath awareness. Both groups—those practicing yoga with and without conscious breath awareness—showed significant reductions in perceived stress levels. However, the difference between the groups was not statistically significant, suggesting that while breath awareness may offer additional benefits, the overall practice of yoga is effective in stress reduction.

These findings align with previous research indicating the efficacy of yoga interventions in mitigating stress among university populations. For instance, Castellote-Caballero et al. reported significant improvements in perceived stress, emotional well-being, and anxiety levels following a 12-week yoga program among university students ^[1]^. Similarly, Lemay et al. observed positive psychophysiological effects of yoga on stress in college students, highlighting its role in stress management ^[17]^.

The marginally greater reduction in stress observed in the breath awareness group may be attributed to the influence of controlled breathing on the autonomic nervous system. Yogic breathing techniques, such as pranayama, have been shown to enhance parasympathetic activity and reduce sympathetic arousal, leading to a state of physiological relaxation. This modulation of the autonomic nervous system can result in decreased cortisol levels and improved stress resilience ^[23]^.

Furthermore, integrating breath awareness into yoga practice may offer cognitive benefits. Qi et al. found that breathing-focused yoga practice led to higher mindfulness and reduced stress compared to movement-focused yoga among undergraduate students ^[23]^. These findings suggest that breath-centric practices may enhance the psychological benefits of yoga by promoting greater present-moment awareness and emotional regulation.

The current study’s findings are consistent with those of other randomized controlled trials. For example, Brandão et al. demonstrated significant reductions in stress and anxiety levels among students following a six-week online Kundalini Yoga program during the COVID-19 pandemic ^[18]^. Additionally, Thakur et al. reported that Heartfulness meditation practice led to decreased anxiety and perceived stress, along with increased well-being and mindfulness among university students ^[24]^.

Moreover, Vollbehr et al. conducted a systematic review and meta-analysis indicating that Hatha yoga is effective in reducing symptoms of mood and anxiety disorders ^[25]^. Ahmad et al. found that both mindfulness and yoga interventions significantly reduced depression and anxiety in college students ^[26]^. Ebert et al.’s meta-analysis further supports the effectiveness of meditation, yoga, and mindfulness in alleviating depression, anxiety, and stress among tertiary education students ^[27]^.

While the present study did not find statistically significant differences between the breath awareness and non-breath awareness groups, the overall reduction in stress levels underscores the therapeutic potential of yoga interventions in academic settings. Given the high prevalence of stress among university students, incorporating structured yoga programs into university wellness initiatives may serve as an effective, non-pharmacological approach to stress management.

Future research should explore the long-term effects of yoga interventions with larger and more diverse samples. Including objective physiological measures, such as heart rate variability and cortisol levels, alongside self-report assessments, could provide a more comprehensive understanding of the mechanisms through which yoga and breath

Awareness influence stress. Additionally, qualitative studies examining students’ experiences with yoga practice may offer insights into factors influencing adherence and perceived benefits.

In conclusion, this study reinforces the growing body of evidence supporting yoga as an effective intervention for reducing stress among university students. While both yoga practices—with and without breath awareness—proved beneficial, the potential added advantages of breath-centric practices warrant further investigation. Integrating yoga programs into university health services could play a pivotal role in enhancing students’ mental well-being and academic performance.

### Limitations

This study has few limitations. The small sample size may have reduced statistical power, potentially contributing to the lack of significant inter-group differences. Short intervention duration (30 days) may not allow sustained effects to manifest, longer programs have shown better outcomes ^[28]^. Reliance on self-reported PSS-10 data limits insight into physiological mechanisms, which could be better understood through biomarkers like cortisol ^[29].^Additionally, lifestyle variables such as sleep, diet, and academic load were not controlled, which may have influenced outcomes. Future research should consider these confounders and individual experiences to improve program efficacy ^[30].^

## Conclusions

This study examined the effects of yoga with and without breath awareness on perceived stress among university students. Using a randomized controlled design, participants were divided into two groups and practiced yoga for 30 days. Stress levels, measured using the Perceived Stress Scale (PSS-10), significantly decreased in both groups post-intervention. Although the group practicing breath awareness showed a slightly greater reduction, the difference was not statistically significant. These results suggest that yoga is effective in reducing stress, with breath awareness potentially offering added benefits. Future studies with larger samples, longer durations, and physiological measures are needed to confirm and expand upon these findings.

## Data Availability

All data produced in the present work are contained in the manuscript.

